# CRISPR/Cas12a-mediated ultrasensitive and on-site monkeypox viral testing

**DOI:** 10.1101/2022.10.10.22280931

**Authors:** Furong Zhao, Pei Wang, Haoxuan Wang, Sirui Liu, Muhammad Sohail, Xing Zhang, Bingzhi Li, He Huang

## Abstract

The unexpected transmission of monkeypox virus (MPXV) from Central and West Africa to previously non-endemic locations is triggering a global panic. The ultrasensitive, rapid, and specific detection of MPXV is crucial for controlling its spreading, while such technology has rarely been reported. Herein, we proposed an MPXV assay combining recombinase-aided amplification (RAA) and CRISPR/Cas12a for the first time. This assay targeted MPXV *F3L* gene and yielded a low detection limit (LOD) of 10^1^ copies/μL. Deriving from the high specificity nature of RAA and CRISPR/Cas12a, through rational optimizations of probes and conditions, this assay showed high selectivity that could distinguish MPXV from other orthopox viruses and current high-profile viruses. To facilitate on-site screening of potential MPXV carriers, a kit integrating lateral flow strips was developed, enabling naked-eye MPXV detection with a LOD of 10^4^ copies/μL. Our RAA-Cas12a-MPXV assay was able to detect MPXV without the need for sophisticated operation and expensive equipment. We envision that this RAA-Cas12a-MPXV assay can be deployed in emerging viral outbreaks for on-site surveillance of MPXV.

**For TOC only:** 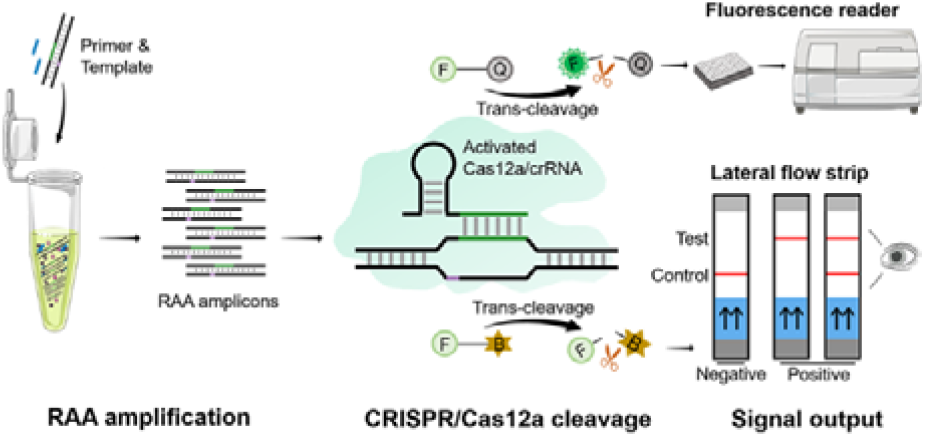

## Introduction

As a member of the genus orthopoxvirus (OPXV) in the family *Poxviridae*, monkeypox virus (MPXV) is a double-stranded DNA (dsDNA) virus firstly isolated in 1958.^1-3^ Monkeypox was not recognized as a zoonotic disease until the first human case discovered in a 9-month-old boy in Zaire (now the Democratic Republic of the Congo) in 1970.^4-6^ Animal-to-human transmission of MPXV can occur via humans’ close contact with infected animals while human-to-human transmission includes contact with bodily fluids, respiratory droplets, and lesion materials of the infected individual, sexual transmission, and maternal-infant transmission.^7, 8^ Since the first human case, monkeypox has become predominantly endemic in Central and West Africa, for example, Nigeria experienced a large monkeypox outbreak in 2017, with relevant cases continuously emerging.^9-11^ It was not until 2003 that the first case occurred in the United States, mainly due to the importation of infected pet prairie dogs from Ghana. Since 2018, monkeypox has also been reported in tourists from Nigeria to other non-endemic countries, involving Israel,^12^ the United Kingdom,^13, 14^ Singapore,^15^ the United States of America,^16, 17^ and others.^18^ Disturbingly, the World Health Organization (WHO) has collected a total of 71,237 laboratory-confirmed cases and 1,097 probable cases from 107 infected member countries since 1 January 2022, and declared this outbreak a public health emergency of international concern.^19^ It has become a new reality that human monkeypox is no longer a rare zoonotic disease, which deserves more public health attention.^20^ Thus, the detection of monkeypox matters even more in the current post-COVID, more vigilant world.^21^

Currently, the MPXV testing relies on methods including polymerase chain reaction (PCR),^22-24^ enzyme-linked immunosorbent assay (ELISA),^25, 26^ loop-mediated isothermal amplification (LAMP),^27^ and microarray.^28, 29^ PCR, recognized as the gold standard for the diagnosis of various diseases including monkeypox, displays high accuracy and sensitivity, yet its application in low-resource areas is limited due to its requirements for expensive equipment and skilled operators.^30^ ELISA may fail in providing MPXV-specific analysis for the serologically cross-reactive characteristic of OPXV and lead to false positive results for recent or remote vaccination.^31^ LAMP is free from precise thermo-cycling, but it suffers from complicated primer design, false positives, and poor quantitative performance. Besides, the microarray method has been developed for the detection and discrimination of OPXV species; however, its practicality and accessibility is limited deriving from high cost and long detection time.^32^ Therefore, low-cost, ultrasensitive, and rapid methods that can facilitate on-site and facile MPXV detection have remained elusive.

CRISPR/Cas (clustered regularly interspaced short palindromic repeats and CRISPR-associated protein) discovered in the adaptive immune system of prokaryotes, was initially proposed as a revolutionary gene editing technology and later expanded to the fields of molecular diagnosis and biosensing.^33, 34^ CRISPR/Cas12a system that integrates biorecognition (Cas12a crRNA binding to the target) with signal transduction (activated Cas12a indiscriminately cleaving any surrounding single-stranded DNA reporters) has participated in the detection of various viruses, including severe acute respiratory syndrome coronavirus 2 (SARS-CoV-2),^35^ African swine fever virus (ASFV),^36^ human immunodeficiency virus (HIV),^37^ displaying obvious advantages of mild conditions, operational simplicity, great sensitivity, and potent signal amplification.^38, 39^ Currently, the CRISPR/Cas12a-based MPXV assay has not been systematically reported. According to the best of our acknowledge, there is only one short letter discussed the possibility of implementing CRISPR technology in MPXV detection, while details on pre-amplification, probe screening, analytical performance, and point-of-care testing (POCT) have not been included.^40^

Herein, we proposed an ultrasensitive and rapid MPXV assay that integrates recombinase-aided amplification (RAA) with CRISPR/Cas12a (the RAA-Cas12a-MPXV assay) for the first time. After RAA pre-amplification of DNA templates (containing MPXV *F3L* gene), CRISPR/Cas12a-based amplicon recognition, and rapid cleavage of reporters, ultrasensitive detection of MPXV down to 10^1^ copies/μL via fluorescence readout was accomplished, which is 1000-fold more sensitive than PCR. For MPXV POCT, we chose lateral flow strip technology as the reliable interpretation of the assay and achieved the detection limit of 10^4^ copies/μL. Our RAA-Cas12a-MPXV assay has the potential to detect MPXV with excellent sensitivity, and selectivity with the aid of basic heaters, providing a rapid, convenient, visual method for on-site screening in low-resource regions.

## Results and Discussion

### Principle and verification of RAA-Cas12a-MPXV Assay

The principle of our RAA-Cas12a-MPXV assay is illustrated in Scheme 1. It consists of three steps: RAA amplification, CRISPR/Cas12a cleavage, and signal output. In the first step, RAA selecting the DNA template, the conserved region of MPXV-specific *F3L* gene,^41-43^ as the target sequence, produces numerous amplicons enhanced the sensitivity of the assay. In the second step, crRNA-specific recognition and binding of amplicons activate the *trans*-cleavage activity of Cas12a, resulting in a large number of ssDNA reporters cleaved. Depending on different reporters, the third step presents two modes of signal output, fluorescence assay for FQ reporters (modified with 6-FAM at the 5’ end and BHQ1 at the 3’ end) and lateral flow strip assay for FB reporters (modified with 6-FAM at the 5’ end and biotin at the 3’ end), improving the suitability and usability of our RAA-Cas12a-MPXV assay. The departure of fluorophores of FQ reporters from quenchers enables fluorescence assay using a fluorescence reader. In lateral flow strip assay, biotin of FB reporters is captured by streptavidin at the first line on the strip (control), while the FAM/anti-FAM antibody/AuNP complex migrates toward the second line (test) and bind to anti-FAM. The result is visible within 5 min after sample addition at room temperature, and the appearance of red color at the test band is considered positive.

**Scheme 1.**
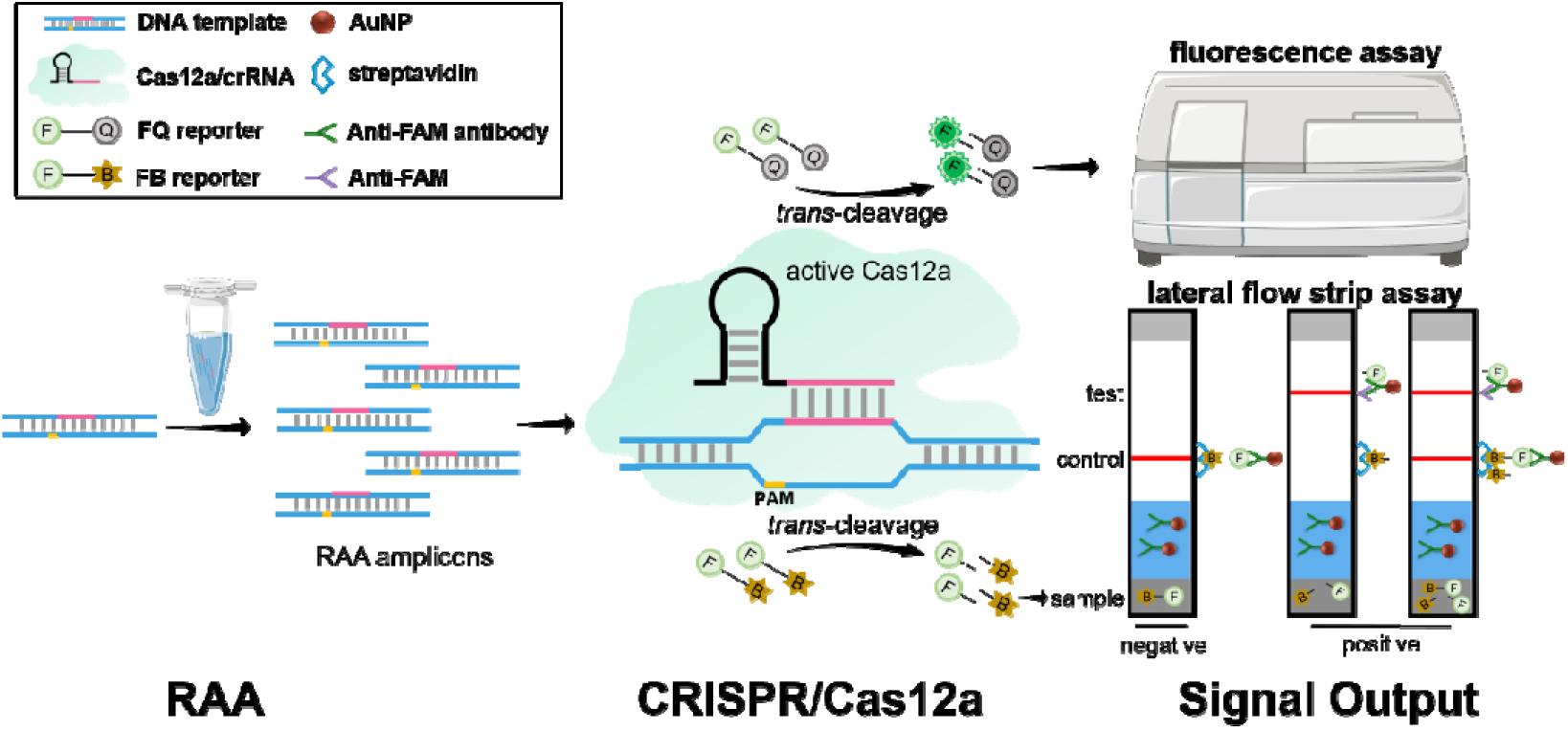
The principle of RAA-Cas12a-MPXV assay proposed in this research. RAA for DNA template amplification; CRISPR/Cas12a system for reporter cleavage; fluorescence assay and lateral flow strip assay for detection signal output.

To assess the feasibility of our RAA-Cas12a-MPXV assay, we compared fluorescence results with or without DNA templates, and RAA product. As shown in Figure 1A, the control group without DNA templates brought no significant fluorescence change (NTC, blue line), indicating that no RAA amplicon was generated for recognition by subsequent CRISPR/Cas12a in the absence of DNA templates. The blank control without RAA product also showed no fluorescent signal, demonstrating that CRISPR/Cas12a system does not cleave reporters without amplicons (NRP, black line). Accordingly, we excluded the possibility of false positive results due to contamination and non-specific amplification during RAA, as well as non-specific cleavage of reporters during CRISPR/Cas12a. In contrast, in the presence of DNA templates, RAA replicated numerous amplicons for crRNA recognition and Cas12a activation, resulting in an intense fluorescence signal (red line). The results obtained by the fluorescence reader were consistent with the fluorescence spectroscopy (Figure 1B). Moreover, the fluorescence signal could be observed by the naked eyes under ultraviolet (UV) light (Figure 1B, inset).

**Figure 1.**
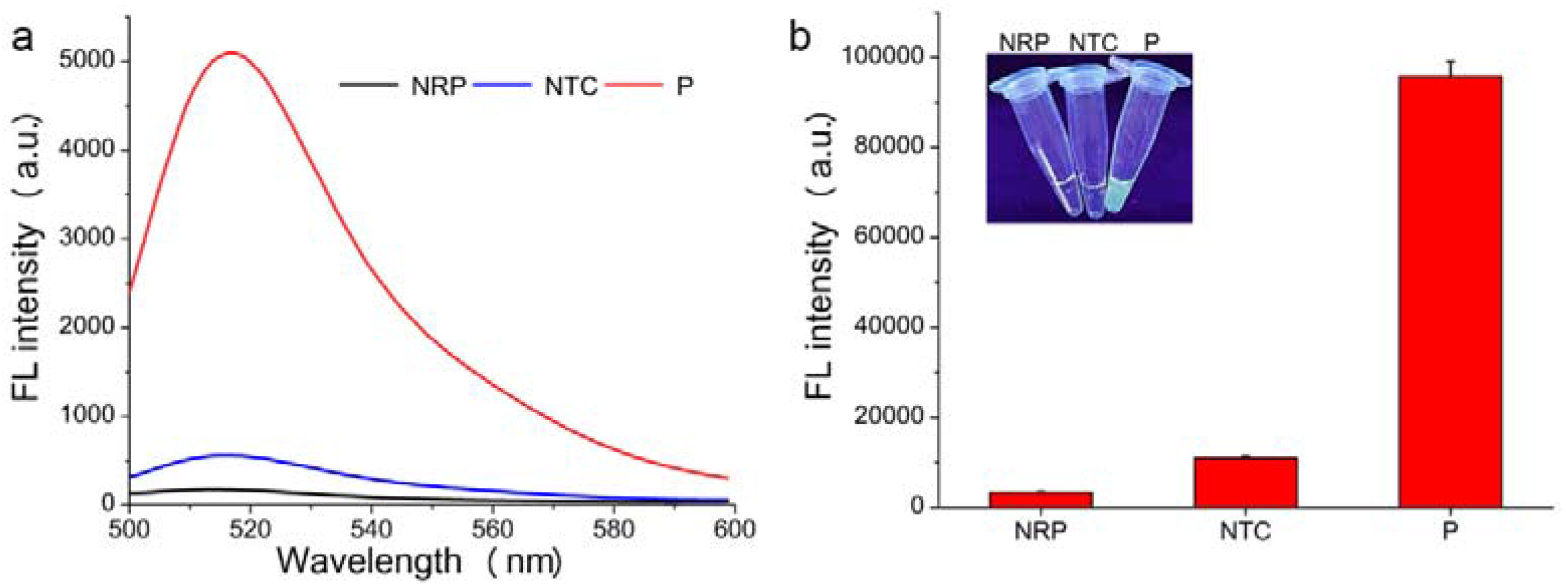
Verification of RAA-Cas12a-MPXV testing strategy. (a) Fluorescence spectra of feasibility verification. (b) Fluorescence intensity of feasibility verification. The inset was photographed by a smartphone under UV light. RAA was carried out under 42 °C for 30 min using F2 and R2, then 1 μL RAA product was added to 20 μL CRISPR/Cas12a reaction (50 nM Cas12a, 50 nM CrRNA, 1 μM FQ reporter, 1 × Cas Buffer). NRP, no RAA product; NTC, no template control; P, positive.

### Design and screening of RAA primers and Cas12a crRNA

RAA primers are the main factors affecting RAA amplification efficiency. To obtain higher sensitivity and selectivity, three pairs of primers binding at different sites of the MPXV-specific *F3L* gene were designed (Figure 2a, Table S1). Agarose gel electrophoresis (AGE) and the RAA-Cas12a-MPXV fluorescence assay were performed to screen the optimal primers for further experiments. The result of AGE suggested that amongst the three pairs of RAA primers, only the third pair (F2+R2) achieved the brightest specific amplification band while having no non-specific amplification band (Figure 2b). The fluorescence assay further confirmed F2 and R2 as the most efficient primers, with the lowest fluorescence in NTC and the highest fluorescence in P (Figure 2c).

**Figure 2.**
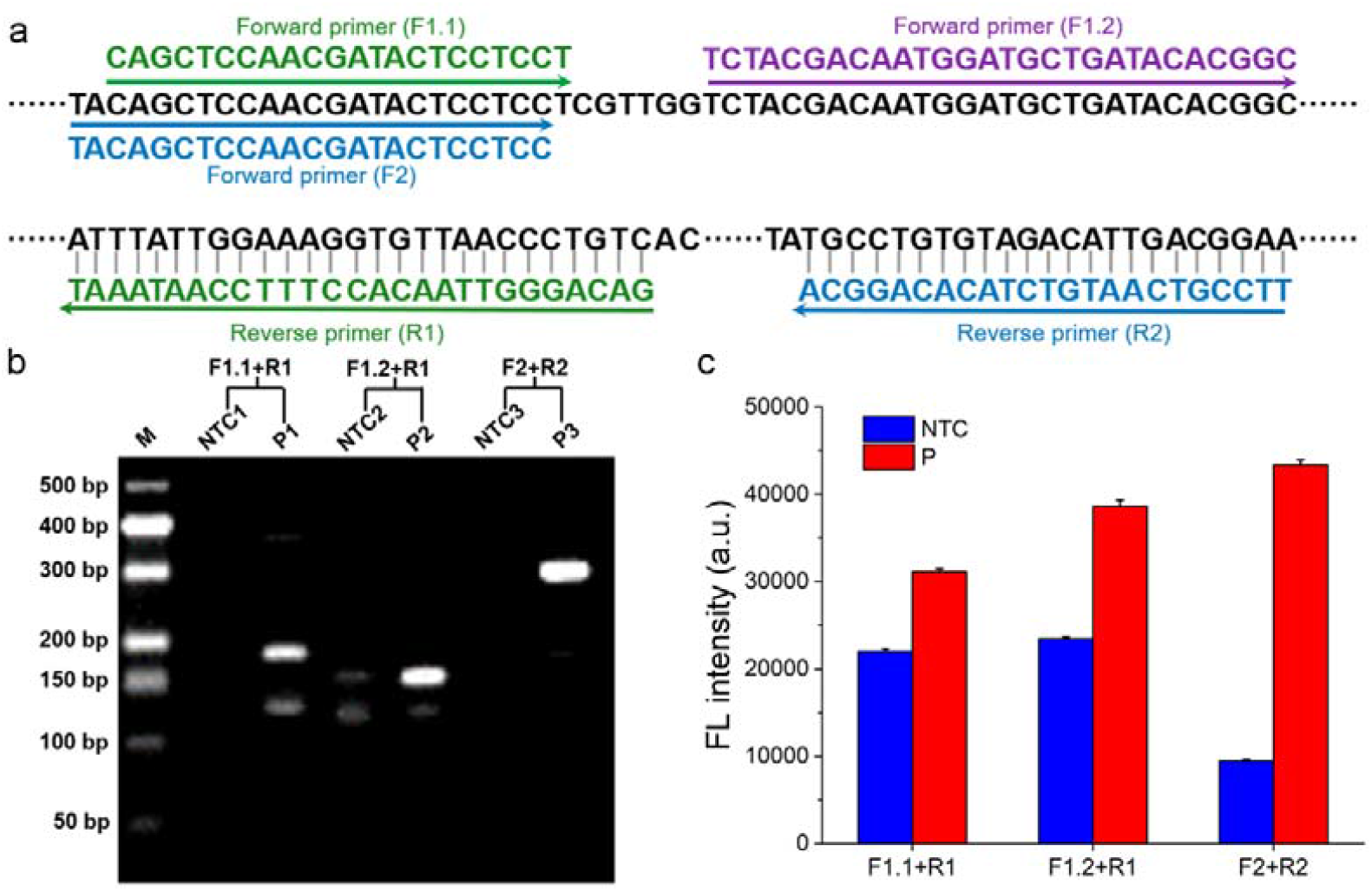
Screening optimal primers for RAA. (a) The binding sites of designed primers on the DNA template. (b) AGE of RAA product. M, DNA marker; lane NTC1 and P1, RAA products using F1.1 and R1; lane NTC2 and P2, RAA products using F1.2 and R1; lane NTC3 and P3, RAA products using F2 and R2. (c) Fluorescence intensity of RAA-CRISPR/Cas12a assay. NTC, no template control; P, positive.

Cas12a crRNA plays a central role in amplicon recognition and Cas12a activation, thus, to obtain the optimized crRNA that can potently activate Cas12a, we designed three crRNAs with the same length, while targeting to different sites on the RAA amplicon (Figure 3a). All of the crRNAs we designed activated Cas12a which subsequently cis-cleaved amplicons (no bands in the insert of Figure 3b) and trans-cleaved FQ reporters (rising fluorescence in Figure 3b), while crRNA2 was the best candidate for its highest fluorescence intensity. With crRNA2-activating CRISPR/Cas12a, we observed a considerable fluorescence difference between the negative group and the positive group (Figure 3c). Consequently, we selected crRNA2 for the following study.

**Figure 3.**
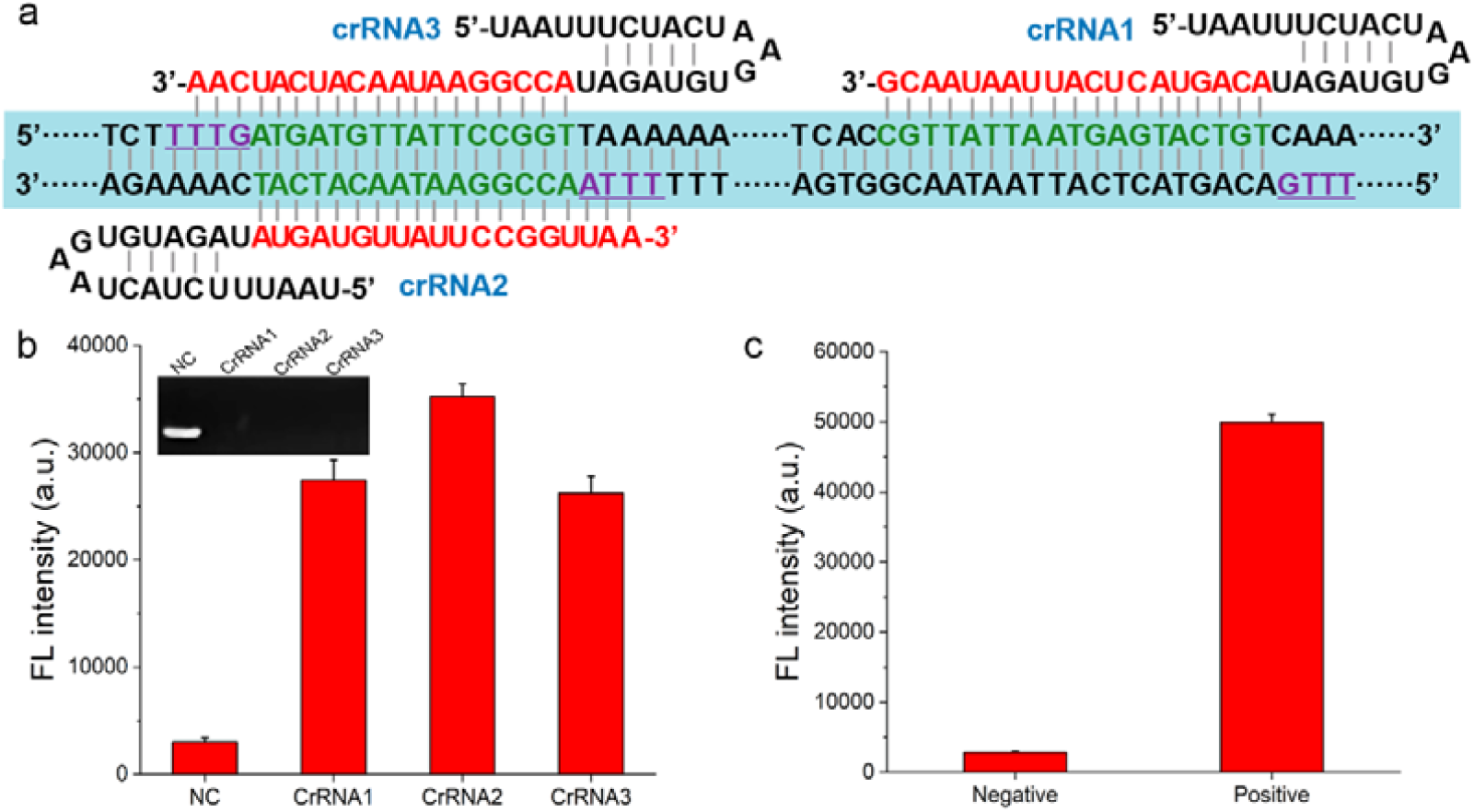
Screening an optimal crRNA for CRISPR/Cas12a. (a) The binding sites of designed crRNAs on the DNA template (Green, crRNA binding sites; purple, PAM motifs). (b) Fluorescence intensity of CRISPR/Cas12a assay using different crRNAs. NC, no crRNA. The inset was an agarose gel image corresponding to different groups. Lane NC, CRISPR/Cas12a reaction products with no crRNA; lane crRNA1, CRISPR/Cas12a reaction products using crRNA1; lane crRNA2, CRISPR/Cas12a reaction products using crRNA2; lane crRNA3, CRISPR/Cas12a reaction products using crRNA3. (c) Fluorescence intensity of CRISPR/Cas12a assay using the optimum crRNA2. Negative, without templates; positive, with templates.

### Optimization of the assay conditions

To obtain optimal analytical performance of our RAA-Cas12a-MPXV assay, several critical experimental conditions during RAA amplification and the CRISPR/Cas12a system were optimized. We first optimized the temperature and reaction time of RAA based on the optimal primers (F2 and R2). The band of 30-min RAA was significantly brighter than that of other amplification times (Figure 4a), and the fluorescence result reconfirmed 30 min as the ideal RAA reaction time (Figure S1). Likewise, we chose 42 □ as the optimal temperature for RAA by combining the results of electrophoresis and fluorescence assay (Figure 4b, Figure S1).

**Figure 4.**
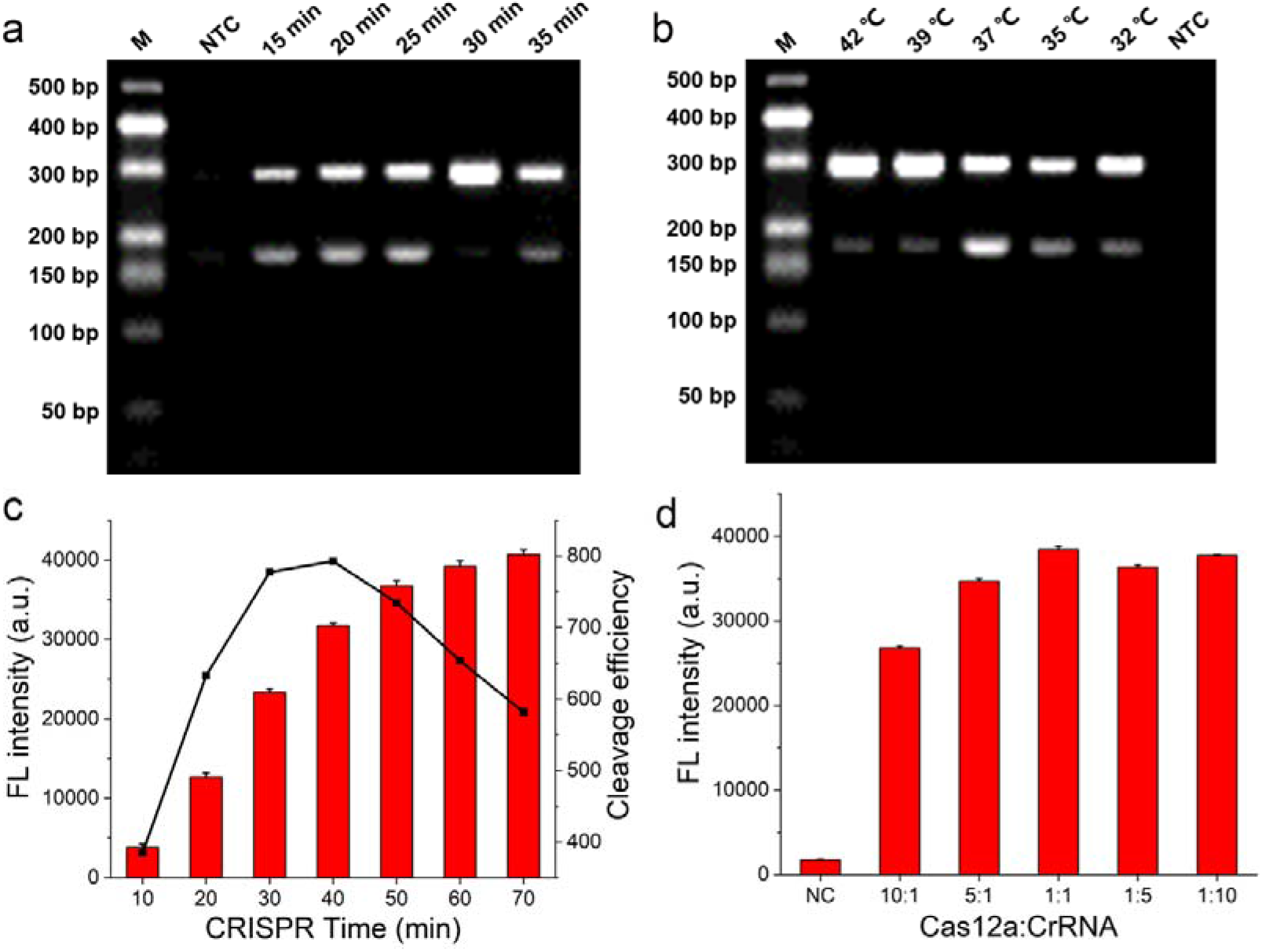
Optimization of the assay conditions. (a) The agarose gel image of RAA amplification products under different reaction times (15 min, 20 min, 25 min, 30 min, 35 min). (b) The agarose gel image of RAA amplification products under different temperatures (42 □, 39 □, 37 □, 35 □, 32 □). NTC, no template control. (c) Fluorescence intensity of CRISPR/Cas12a assay under different reaction times (10 min, 20 min, 30 min, 40 min, 50 min, 60 min, 70 min). Cleavage efficiency = fluorescence intensity/CRISPR time. (d) Fluorescence intensity of CRISPR/Cas12a assay using different ratios of Cas12a and crRNA (10:1, 5:1, 1:1, 1:5, 1:10). NC, no crRNA.

After the determination of RAA amplification conditions (42 □, 30 min), we further optimized the CRISPR/Cas12a system with crRNA2. We first investigated the cleavage ability of Cas12a at different reaction times. The fluorescence intensity spiked rapidly from the onset of the reaction to 40 min, and the cleavage efficiency reached its maximum at 40 min, after which the fluorescence slowly increased (Figure 4c), thus, we chose 40 min as the best CRISPR reaction time. Furthermore, we explored the effects of Cas12a:crRNA on the ultimate fluorescence intensity and found that 1:1 was the optimal ratio (Figure 4d).

### Performances of the proposed RAA-Cas12a-MPXV fluorescence assay

As shown in Figure 5a, our proposed RAA-Cas12a-MPXV fluorescence assay has the merits of easy operation and rapid testing. To evaluate the sensitivity of the fluorescence assay, DNA templates at serial dilution concentrations in the range of 10^0^ to 10^10^ copies/μL were subjected to RAA amplification, and then a 1 μL amplification product was taken for the CRISPR/Cas12a reaction. The resulting fluorescence intensity in Figure 5a exhibited that compared to the control group without the DNA template and RAA product, the difference initiated in the group of 10^1^ copies/μL, and gradually widened in the following groups, hence the limit of detection (LOD) of the RAA-Cas12a-MPXV fluorescence assay was 10^1^ copies/μL. Additionally, we achieved the LOD of 10^2^ copies/μL via naked-eye observation under UV light (Figure 5b, insert). We also evaluated the sensitivity of the PCR-Cas12a-MPXV fluorescence assay (detailed in Supporting Information). In comparison with the PCR-Cas12a-MPXV fluorescence assay, the LOD of which was 10^4^ copies/μL (Figure S2), the RAA-Cas12a-MPXV fluorescence assay we established achieved 1000-fold lower LOD, performing outstanding sensitivity. The higher sensitivity of RAA-based assay is ascribed to the higher amplification efficiency of RAA and the better compatibility of RAA to CRISPR.^44-46^

**Figure 5.**
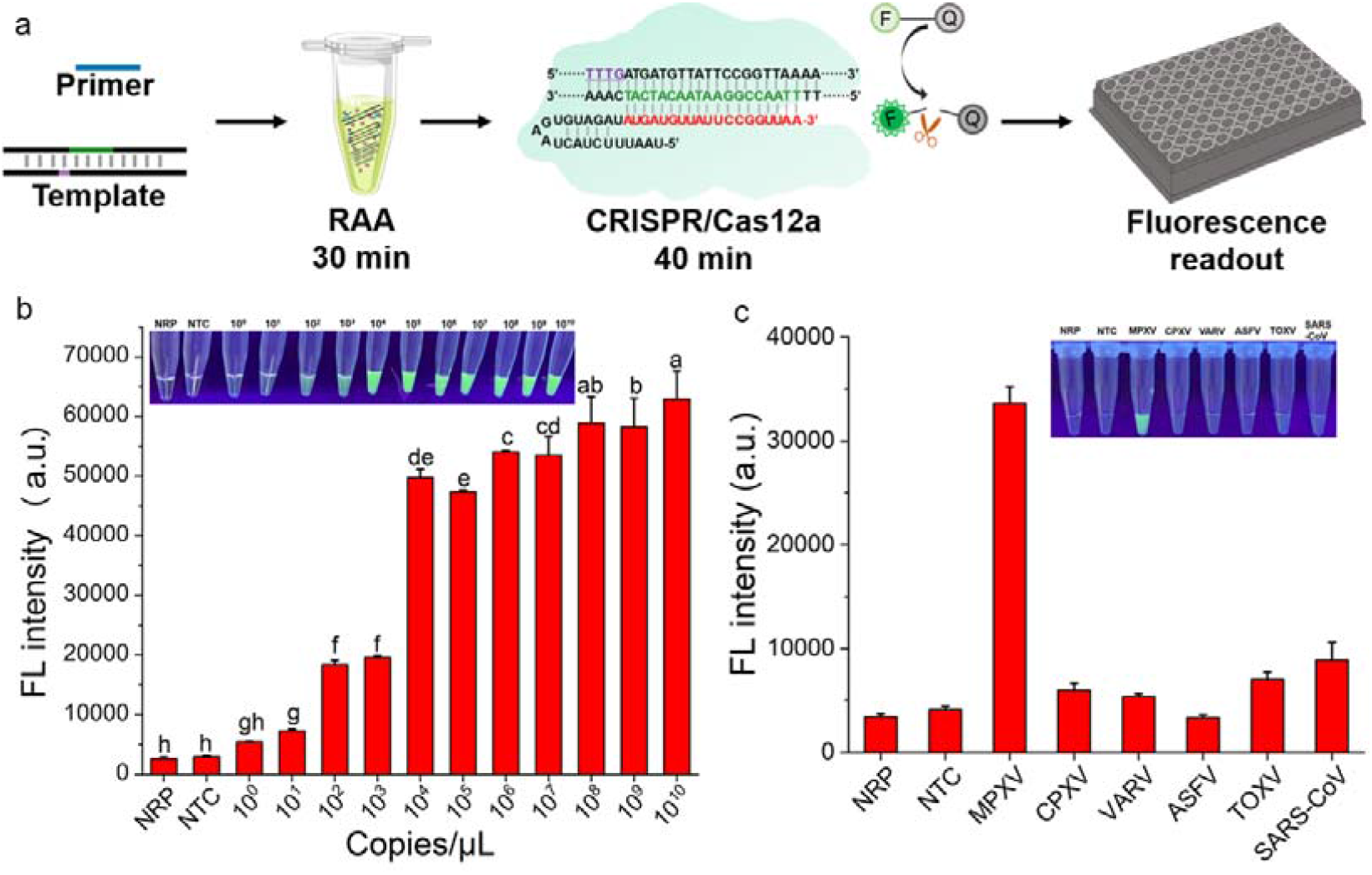
Performances of the RAA-Cas12a-MPXV fluorescence assay. (a) Flowchart of the fluorescence assay. (b) Fluorescence intensity responding to a serial 10-fold dilution (10^10-0^ copies/μL) of DNA templates in the sensitivity evaluation. The inset was photographed by a smartphone under UV light. Copy numbers and serial dilution values that are significantly different from each other are labeled with different lowercase letters above the error bars (p < 0.05). (c) Fluorescence intensity responding to MPXV, CPXV, VARV, ASFV, TOXV, and SARA-CoV-2 in the selectivity evaluation. The inset was photographed by a smartphone under UV light. NRP, no RAA product; NTC, no template control.

The selectivity of our RAA-Cas12a-MPXV fluorescence assay was next assessed by comparing the fluorescence intensity produced by MPXV with that produced by other viruses: cowpox virus (CPXV), variola virus (VARV), ASFV, and *Toxoplasma Gondii* virus (TOXV), SARS-CoV-2. Both fluorescence values and naked-eye observation demonstrated that only the MPXV group induced enhanced fluorescence while other orthopox viruses (CPXV and VARV), and current high-profile viruses (ASFV, TOXV, SARS-CoV-2) showed no fluorescence changes (Figure 5c). These results confirmed the superior selectivity of our strategy toward MPXV, which is derived from the outstanding specificity of RAA amplification and Cas12a recognition.^47-49^

### RAA-Cas12a-MPXV Lateral Flow Strip Assay for Point-of-Care Testing

To enable POCT, we designed the FB reporter as a substitute for the FQ reporter of the fluorescence assay and established the RAA-Cas12a-MPXV lateral flow strip assay (Figure 6a). Upon the addition of the reaction solution containing both intact FB reporters and cracked FAMs and biotins at the sample pad, the anti-FAM antibody/AuNP complexes quickly bound to the isolated FAMs as well as the intact FB reporters as the sample migrated forward. Subsequently, the streptavidin, which was immobilized at the control band, captured the cracked biotins and the FB reporter/anti-FAM antibody/AuNP complexes, so that the control band appeared red owing to the aggregation of AuNPs. The rest isolated FAM/anti-FAM antibody/AuNP complexes migrated toward the test band and were captured by the FAM antibodies, appearing red. Once there were enough amplicons to activate Cas12a to cleave all FB reporters at maximum capacity, the color change would only appear at the test band as a result. In brief, the color change appearing at the test band only or both the control band and the test band indicated a positive result. In the negative group, the absence of the DNA templates led to no amplicons for Cas12a activation and reporters cleavage, consequently, the red color appeared only at the control band.

**Figure 6.**
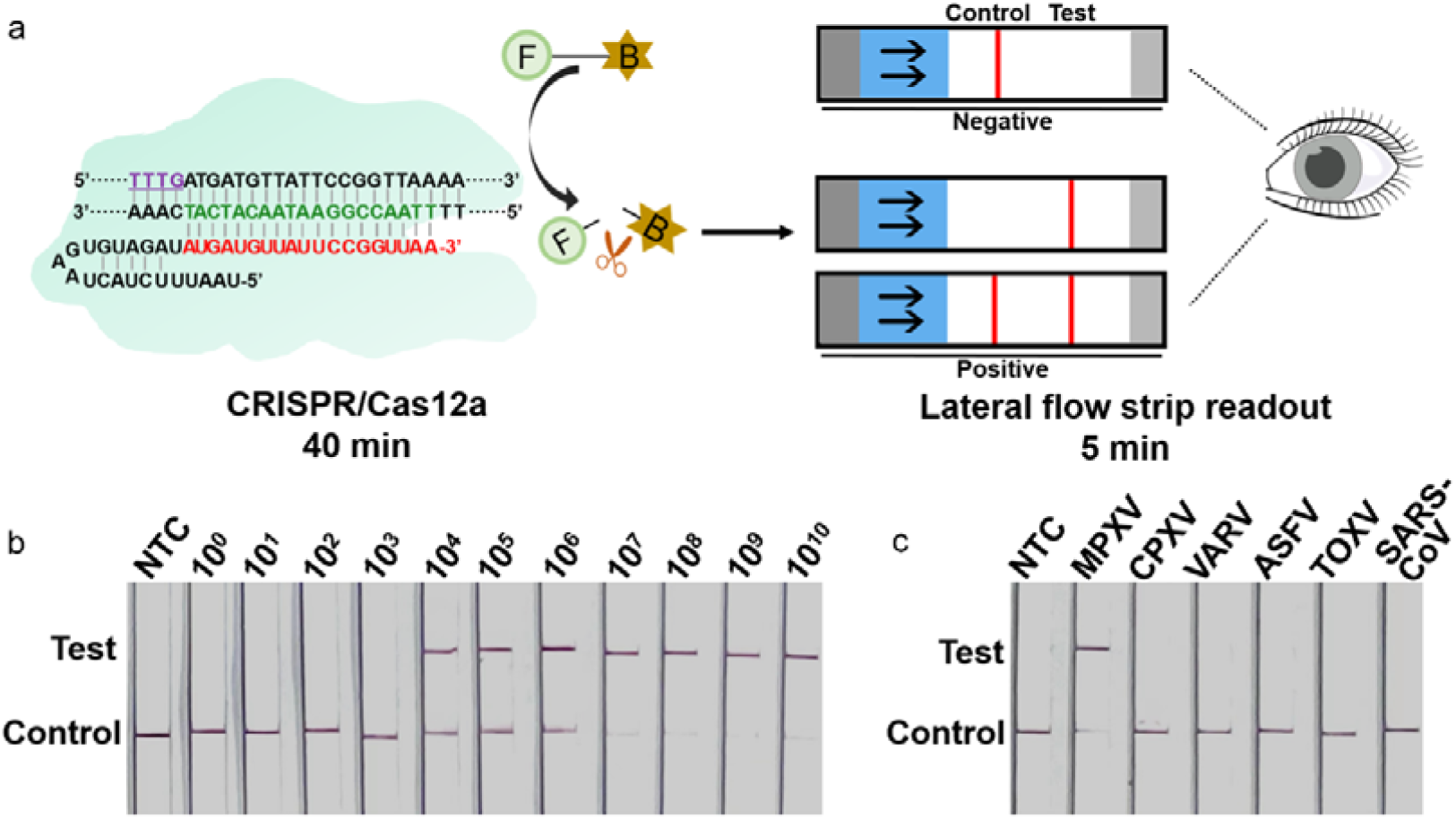
Performances of the RAA-Cas12a-MPXV lateral flow strip assay. (a) Flowchart of the lateral flow strip assay. (b) Lateral flow strip results responding to a serial 10-fold dilution (10^10-0^ copies/μL) of DNA templates in the sensitivity evaluation. (c) Lateral flow strip results responding to MPXV, CPXV, VARV, ASFV, TOXV, and SARA-CoV-2 in the selectivity evaluation. NTC, no template control.

In the sensitivity assessment, with the increasing number of DNA templates, amplicons for crRNA recognition and Cas12a activation incrementally grew via RAA, eventually leading to a growing number of FB reporters being cleaved into FAM and biotin, allowing the result that the color became darker at the test band and became lighter at the control band presented on the lateral flow strips. Through naked-eye observation, our RAA-Cas12a-MPXV lateral flow strip assay achieved the LOD of 104 copies/μL (Figure 6b). Besides, this lateral flow strip assay showed great selectivity, as demonstrated by the cross-reactivity test with other orthopox viruses and current high-profile viruses (Figure 6c).

## Conclusions

In this work, we established an RAA-Cas12a-MPXV assay for ultrasensitive, facile, and rapid detection of MPXV by integrating RAA amplification with CRISPR/Cas12a system and outputting the detection signal in the form of fluorescence or lateral flow strip. Compared to current methods reported for detecting MPXV, our RAA-Cas12a-MPXV assay possessed its notable strengths: (1) our assay was carried out at a gentle temperature throughout, getting rid of the restraints of thermal cycling processes and expensive instruments; (2) compared to the conventional PCR with the LOD of 10^4^ copies/μL, our assay exhibited a significantly improved detection limit of 10^1^ copies/μL in the fluorescence assay, 1000-fold more sensitive than PCR. Meanwhile, visualized detection was realized with the LOD of 10^2^ copies/μL via UV light, and 10^4^ copies/μL via lateral flow strip; (3) combining the lateral flow strip enabled us to perform on-site detection via naked eyes within 5 min at room temperature with no need for any lab equipment, revealing the POCT potential for screening MPXV in low-equipment scenario. Overall, the RAA-Cas12a-MPXV assay is a promising method for MPXV detection with outstanding sensitivity, selectivity, and portability.

## Experimental section

### Materials and Reagents

The oligonucleotides used in this study were synthesized and purified by Sangon Biotech (Shanghai, China) and listed in Table S1. The monkeypox virus *F3L* plasmid was purchased from Sangon Biotech. The RAA nucleic acid amplification kit (S001ZC) was purchased from ZC Bioscience (Hangzhou, China). The 2 × Taq Master Mix (P111) for PCR and the fastpure gel DNA extraction mini kit (DC301) were provided from Vazyme Biotech (Nanjing, China). The Cas12a nuclease was expressed and purified by our laboratory, details were provided in the supporting information. Reagents in the 10 × Cas Buffer (500 mM NaCl, 100 mM Tris-HCl, 100 mM MgCl_2_, 1 mg/mL BSA, pH 7.9) were purchased from Aladdin (Shanghai, China). The lateral flow strip was purchased from IVD Biotech Service Center (Shanghai, China).

### Primers and crRNAs Design

The MPXV *F3L* gene sequence in this study was derived from German strain MPXV-BY-IMB25241 (GenBank: ON568298) in May 2022 and American strain MPXV-USA-2022-MA001 (GenBank: ON563414) during the same period, the difference of which from Zaire strain Zaire-96-l-16 (NCBI Gene ID: 928998) was a partial site mutation. Referring to the previously reported PCR primers for Zaire strains with some changes,^50^ we designed three pairs of RAA primers and screened out the best primers in the follow-up. Cas12a crRNA consisted of a repeat sequence and a amplicon -specific sequence. We designed three 19-nt crRNAs without the PAM motif (TTTN), while the PAM motif appeared at the 5’ end of the non-complementary DNA sequence being targeted.

### Plasmid Construction and Amplification

The gene sequences of CPXV, VARV, ASFV, and *Toxoplasma Gondii* virus (TOXV) were retrieved from GenBank under accession number X75158, AY552594, MN809121 and AF146527, then cloned into PUC57 vectors (Sangon, China) to create the recombinant plasmids. After transformation into *E. coli* DH5α and expansion overnight, the recombinant plasmids were extracted using AxyPrep plasmid DNA mini kit (AP-MN-P-50) purchased from Axygen Biotech (Hangzhou, China). The concentrations of extracted plasmids were determined with NanoDrop 2000 (Thermo Fisher, United States).

### Recombinase-Aided Amplification Reaction

RAA reaction was performed with the RAA nucleic acid amplification kit, following the manufacturer’s instructions. Explicitly, the reaction powder was dissolved with a mixture containing 5 μL A Buffer (provided in the kit), 0.4 μL each of 10 μM forward primer and reverse primer, and 2.7 μL ddH_2_O. Then, 1μL DNA template and 0.5μL B Buffer (provided in the kit) were added into the reaction tube and mixed gently, followed by incubation at 42 °C for 30 min. The DNA template was replaced by ddH_2_O in no template control (NTC) reaction. RAA products were verified by 3 % AGE after purification by the fastpure gel DNA extraction mini kit. The optimum primers for RAA were screened under the observation of the brightness of the specific bands and the absence of non-specific bands after electrophoresis.

### RAA-Cas12a-MPXV Fluorescence Assay

The RAA-Cas12a-MPXV fluorescence assay was performed in a 20 μL system containing 1 × Cas Buffer, 50 nM crRNA, 50 nM Cas12a, 1 μM FQ reporter, and 1 μL RAA product. Cas12a and crRNA were pre-conjugated in Cas Buffer at 37 °C for 10 min to facilitate cleavage performance of Cas12a. After the addition of FQ and RAA product, the CRISPR reaction was incubated at 37 °C for 40 min and observed under the FAM channel of the BioTek Synergy H1M microplate reader (Bio-Tek, United States). In no RAA product (NRP) reaction, ddH_2_O was in place of the RAA product.

### RAA-Cas12a-MPXV Lateral Flow Strip Assay

The reaction system of the RAA-Cas12a-MPXV lateral flow strip assay was similar to the fluorescence assay above, except for the replacement of the FQ with the FB reporter. After incubation at 37 °C for 40 min, sterile ddH_2_O was added to make up the reaction system to 50 μL for the insertion of the lateral flow strip. The result was visualized after 5 min incubation at room temperature. The color appeared only at the control band, indicating a negative result, and the color appeared at the test band or both bands, indicating a positive result.

### Statistical Analysis

Software OriginPro 2021 (OriginLab, Northampton, MA) was used for statistical analyses and graphs. Data were shown as the average ± standard deviation (n = 3). Error bars represent the standard deviation from three independent experiments.

## Data Availability

All data produced in the present study are available upon reasonable request to the authors

## Acknowledgments

This work was supported by grants from the Natural Science Foundation of Jiangsu Province (No. BK20200718 to B.Li), the Natural Science Foundation of the Jiangsu Higher Education Institutions of China (20KJB350010 to B.Li), Jiangsu Innovation and Entrepreneurship Doctoral Program to X.Zhang, and Jiangsu Specially-Appointed Professor Project to X.Zhang.

## References

(1) Xiang, Y.; White, A. Monkeypox virus emerges from the shadow of its more infamous cousin: family biology matters. Emerg Microbes Infec 2022, 11 (1), 1768–1777.

(2) Bunge, E. M.; Hoet, B.; Chen, L.; Lienert, F.; Weidenthaler, H.; Baer, L. R.; Steffen, R. The changing epidemiology of human monkeypox-A potential threat? A systematic review. PLoS Negl Trop Dis 2022, 16 (2), e0010141.

(3) Organization, W. H. Monkeypox. 2022. https://www.who.int/news-room/fact-sheets/detail/monkeypox.

(4) Lai, C. C.; Hsu, C. K.; Yen, M. Y.; Lee, P. I.; Ko, W. C.; Hsueh, P. R. Monkeypox: An emerging global threat during the COVID-19 pandemic. J Microbiol Immunol Infect 2022. DOI: 10.1016/j.jmii.2022.07.004.

(5) McFadden, G. Poxvirus tropism. Nat Rev Microbiol 2005, 3 (3), 201–213.

(6) Plowright, R. K.; Parrish, C. R.; McCallum, H.; Hudson, P. J.; Ko, A. I.; Graham, A. L.; Lloyd-Smith, J. O. Pathways to zoonotic spillover. Nat Rev Microbiol 2017, 15 (8), 502–510.

(7) Murphy, H.; Ly, H. The potential risks posed by inter- and intraspecies transmissions of monkeypox virus. Virulence 2022, 13 (1), 1681–1683.

(8) Di Giulio, D. B.; Eckburg, P. B. Human monkeypox: an emerging zoonosis. Lancet Infect Dis 2004, 4 (1), 15–25.

(9) Reed, K. D.; Melski, J. W.; Graham, M. B.; Regnery, R. L.; Sotir, M. J.; Wegner, M. V.; Kazmierczak, J. J.; Stratman, E. J.; Li, Y.; Fairley, J. A.; et al. The detection of monkeypox in humans in the Western Hemisphere. N Engl J Med 2004, 350 (4), 342–350.

(10) Rimoin, A. W.; Mulembakani, P. M.; Johnston, S. C.; Lloyd Smith, J. O.; Kisalu, N. K.; Kinkela, T. L.; Blumberg, S.; Thomassen, H. A.; Pike, B. L.; Fair, J. N.; et al. Major increase in human monkeypox incidence 30 years after smallpox vaccination campaigns cease in the Democratic Republic of Congo. Proc Natl Acad Sci U S A 2010, 107 (37), 16262–16267.

(11) Durski, K. N.; McCollum, A. M.; Nakazawa, Y.; Petersen, B. W.; Reynolds, M. G.; Briand, S.; Djingarey, M. H.; Olson, V.; Damon, I. K.; Khalakdina, A. Emergence of Monkeypox - West and Central Africa, 1970-2017. MMWR Morb Mortal Wkly Rep 2018, 67 (10), 306–310.

(12) Erez, N.; Achdout, H.; Milrot, E.; Schwartz, Y.; Wiener-Well, Y.; Paran, N.; Politi, B.; Tamir, H.; Israely, T.; Weiss, S.; et al. Diagnosis of Imported Monkeypox, Israel, 2018. Emerg Infect Dis 2019, 25 (5), 980–983.

(13) Hobson, G.; Adamson, J.; Adler, H.; Firth, R.; Gould, S.; Houlihan, C.; Johnson, C.; Porter, D.; Rampling, T.; Ratcliffe, L.; et al. Family cluster of three cases of monkeypox imported from Nigeria to the United Kingdom, May 2021. Euro Surveill 2021, 26 (32), 2100745.

(14) Vaughan, A.; Aarons, E.; Astbury, J.; Balasegaram, S.; Beadsworth, M.; Beck, C. R.; Chand, M.; O’Connor, C.; Dunning, J.; Ghebrehewet, S.; et al. Two cases of monkeypox imported to the United Kingdom, September 2018. Euro Surveill 2018, 23 (38), 2–6.

(15) Ng, O. T.; Lee, V.; Marimuthu, K.; Vasoo, S.; Chan, G. H.; Lin, R. T. P.; Leo, Y. S. A case of imported Monkeypox in Singapore. Lancet Infect Dis 2019, 19 (11), 1166–1166.

(16) Rao, A. K.; Schulte, J.; Chen, T. H.; Hughes, C. M.; Davidson, W.; Neff, J. M.; Markarian, M.; Delea, K. C.; Wada, S.; Liddell, A.; et al. Monkeypox in a Traveler Returning from Nigeria - Dallas, Texas, July 2021. Mmwr-Morbid Mortal W 2022, 71 (14), 509–516.

(17) Costello, V.; Sowash, M.; Gaur, A.; Cardis, M.; Pasieka, H.; Wortmann, G.; Ramdeen, S. Imported Monkeypox from International Traveler, Maryland, USA, 2021 (Response). Emerg Infect Dis 2022, 28 (8), 1738–1738.

(18) Yinka-Ogunleye, A.; Aruna, O.; Dalhat, M.; Ogoina, D.; McCollum, A.; Disu, Y.; Mamadu, I.; Akinpelu, A.; Ahmad, A.; Burga, J.; et al. Outbreak of human monkeypox in Nigeria in 2017-18: a clinical and epidemiological report. Lancet Infect Dis 2019, 19 (8), 872–879.

(19) Organization, W. H. 2022 Monkeypox Outbreak: Global Trends. 2022. https://worldhealthorg.shinyapps.io/mpx_global/.

(20) Huhn, G. D.; Bauer, A. M.; Yorita, K.; Graham, M. B.; Sejvar, J.; Likos, A.; Damon, I. K.; Reynolds, M. G.; Kuehnert, M. J. Clinical characteristics of human monkeypox, and risk factors for severe disease. Clin Infect Dis 2005, 41 (12), 1742–1751.

(21) Dzobo, M.; Gwinji, P. T.; Murewanhema, G.; Musuka, G.; Dzinamarira, T. Stigma and public health responses: Lessons learnt from the COVID-19 pandemic to inform the recent monkeypox outbreak. Public Health Pract (Oxf) 2022, 4, 100315.

(22) Li, Y.; Zhao, H.; Wilkins, K.; Hughes, C.; Damon, I. K. Real-time PCR assays for the specific detection of monkeypox virus West African and Congo Basin strain DNA. J Virol Methods 2010, 169 (1), 223–227.

(23) Chelsky, Z. L.; Dittmann, D.; Blanke, T.; Chang, M.; Vormittag-Nocito, E.; Jennings, L. J. Validation Study of a Direct Real-Time PCR Protocol for Detection of Monkeypox Virus. J Mol Diagn 2022. DOI: 10.1016/j.jmoldx.2022.09.001.

(24) Huggett, J. F.; French, D.; O’Sullivan, D. M.; Moran-Gilad, J.; Zumla, A. Monkeypox: another test for PCR. Euro Surveill 2022, 27 (32).

(25) Karem, K. L.; Reynolds, M.; Braden, Z.; Lou, G.; Bernard, N.; Patton, J.; Damon, I. K. characterization of acute-phase humoral immunity to monkeypox: use of immunoglobulin M enzyme-linked immunosorbent assay for detection of monkeypox infection during the 2003 North American outbreak. Clin Diagn Lab Immunol 2005, 12 (7), 867–872.

(26) Dubois, M. E.; Hammarlund, E.; Slifka, M. K. Optimization of peptide-based ELISA for serological diagnostics: a retrospective study of human monkeypox infection. Vector Borne Zoonotic Dis 2012, 12 (5), 400–409.

(27) Iizuka, I.; Saijo, M.; Shiota, T.; Ami, Y.; Suzaki, Y.; Nagata, N.; Hasegawa, H.; Sakai, K.; Fukushi, S.; Mizutani, T.; et al. Loop-mediated isothermal amplification-based diagnostic assay for monkeypox virus infections. J Med Virol 2009, 81 (6), 1102–1108.

(28) Ryabinin, V. A.; Shundrin, L. A.; Kostina, E. B.; Laassri, M.; Chizhikov, V.; Shchelkunov, S. N.; Chumakov, K.; Sinyakov, A. N. Microarray assay for detection and discrimination of Orthopoxvirus species. J Med Virol 2006, 78 (10), 1325–1340.

(29) Kostina, E. V.; Sinyakov, A. N.; Ryabinin, V. A. A many probes-one spot hybridization oligonucleotide microarray. Anal Bioanal Chem 2018, 410 (23), 5817–5823.

(30) Altindis, M.; Puca, E.; Shapo, L. Diagnosis of monkeypox virus - An overview. Travel Med Infect Dis 2022, 50, 102459.

(31) Edghill-Smith, Y.; Golding, H.; Manischewitz, J.; King, L. R.; Scott, D.; Bray, M.; Nalca, A.; Hooper, J. W.; Whitehouse, C. A.; Schmitz, J. E.; et al. Smallpox vaccine-induced antibodies are necessary and sufficient for protection against monkeypox virus. Nat Med 2005, 11 (7), 740–747.

(32) Berthet, N.; Dickinson, P.; Filliol, I.; Reinhardt, A. K.; Batejat, C.; Vallaeys, T.; Kong, K. A.; Davies, C.; Lee, W.; Zhang, S.; et al. Massively parallel pathogen identification using high-density microarrays. Microb Biotechnol 2008, 1 (1), 79–86.

(33) Li, S. Y.; Cheng, Q. X.; Wang, J. M.; Li, X. Y.; Zhang, Z. L.; Gao, S.; Cao, R. B.; Zhao, G. P.; Wang, J. CRISPR-Cas12a-assisted nucleic acid detection. Cell Discov 2019, 5 (1), 1.

(34) Xiong, Y.; Zhang, J.; Yang, Z.; Mou, Q.; Ma, Y.; Xiong, Y.; Lu, Y. Functional DNA Regulated CRISPR-Cas12a Sensors for Point-of-Care Diagnostics of Non-Nucleic-Acid Targets. J Am Chem Soc 2020, 142 (1), 207–213.

(35) Xie, S.; Qin, C.; Zhao, F.; Shang, Z.; Wang, P.; Sohail, M.; Zhang, X.; Li, B. A DNA-Cu nanocluster and exonuclease I integrated label-free reporting system for CRISPR/Cas12a-based SARS-CoV-2 detection with minimized background signals. J Mater Chem B 2022, 10 (32), 6107–6117.

(36) Xie, S.; Tao, D.; Fu, Y.; Xu, B.; Tang, Y.; Steinaa, L.; Hemmink, J. D.; Pan, W.; Huang, X.; Nie, X.; et al. Rapid Visual CRISPR Assay: A Naked-Eye Colorimetric Detection Method for Nucleic Acids Based on CRISPR/Cas12a and a Convolutional Neural Network. Acs Synth Biol 2022, 11 (1), 383–396.

(37) Pang, Y.; Li, Q.; Wang, C.; Zhen, S.; Sun, Z.; Xiao, R. CRISPR-cas12a mediated SERS lateral flow assay for amplification-free detection of double-stranded DNA and single-base mutation. Chem Eng J 2022, 429, 132109.

(38) Chen, J. S.; Ma, E.; Harrington, L. B.; Da Costa, M.; Tian, X.; Palefsky, J. M.; Doudna, J. A. CRISPR-Cas12a target binding unleashes indiscriminate single-stranded DNase activity. Science 2018, 360 (6387), 436–439.

(39) Liang, M.; Li, Z.; Wang, W.; Liu, J.; Liu, L.; Zhu, G.; Karthik, L.; Wang, M.; Wang, K. F.; Wang, Z.; et al. A CRISPR-Cas12a-derived biosensing platform for the highly sensitive detection of diverse small molecules. Nat Commun 2019, 10 (1), 3672.

(40) Sui, Y.; Xu, Q.; Liu, M.; Zuo, K.; Liu, X.; Liu, J. CRISPR-Cas12a-based detection of monkeypox virus. J Infect 2022. DOI: 10.1016/j.jinf.2022.08.043.

(41) Kim, J. W.; Lee, M.; Shin, H.; Choi, C. H.; Choi, M. M.; Kim, J. W.; Yi, H.; Yoo, C. K.; Rhie, G. E. Isolation and identification of monkeypox virus MPXV-ROK-P1-2022 from the first case in the Republic of Korea. Osong Public Health Res Perspect 2022, 13 (4), 308–311.

(42) Gong, Q.; Wang, C.; Chuai, X.; Chiu, S. Monkeypox virus: a re-emergent threat to humans. Virol Sin 2022, 37 (4), 477–482.

(43) Feng, J.; Xue, G.; Cui, X.; Du, B.; Feng, Y.; Cui, J.; Zhao, H.; Gan, L.; Fan, Z.; Fu, T.; et al. Development of a Loop-Mediated Isothermal Amplification Method for Rapid and Visual Detection of Monkeypox Virus. Microbiol Spectr 2022, e0271422.

(44) Zhang, Y.; Li, Q.; Guo, J.; Li, D.; Wang, L.; Wang, X.; Xing, G.; Deng, R.; Zhang, G. An Isothermal Molecular Point of Care Testing for African Swine Fever Virus Using Recombinase-Aided Amplification and Lateral Flow Assay Without the Need to Extract Nucleic Acids in Blood. Front Cell Infect Microbiol 2021, 11, 633763.

(45) Ma, Q. N.; Wang, M.; Zheng, L. B.; Lin, Z. Q.; Ehsan, M.; Xiao, X. X.; Zhu, X. Q. RAA-Cas12a-Tg: A Nucleic Acid Detection System for Toxoplasma gondii Based on CRISPR-Cas12a Combined with Recombinase-Aided Amplification (RAA). Microorganisms 2021, 9 (8), 1644.

(46) Zhang, A.; Sun, B.; Zhang, J.; Cheng, C.; Zhou, J.; Niu, F.; Luo, Z.; Yu, L.; Yu, C.; Dai, Y.; et al. CRISPR/Cas12a Coupled With Recombinase Polymerase Amplification for Sensitive and Specific Detection of Aphelenchoides besseyi. Front Bioeng Biotechnol 2022, 10, 912959.

(47) Li, F.; Ye, Q.; Chen, M.; Zhou, B.; Zhang, J.; Pang, R.; Xue, L.; Wang, J.; Zeng, H.; Wu, S.; et al. An ultrasensitive CRISPR/Cas12a based electrochemical biosensor for Listeria monocytogenes detection. Biosens Bioelectron 2021, 179, 113073.

(48) Li, F.; Ye, Q.; Chen, M.; Xiang, X.; Zhang, J.; Pang, R.; Xue, L.; Wang, J.; Gu, Q.; Lei, T.; et al. Cas12aFDet: A CRISPR/Cas12a-based fluorescence platform for sensitive and specific detection of Listeria monocytogenes serotype 4c. Anal Chim Acta 2021, 1151, 338248.

(49) Zhi, S.; Shen, J.; Li, X.; Jiang, Y.; Xue, J.; Fang, T.; Xu, J.; Wang, X.; Cao, Y.; Yang, D.; et al. Development of Recombinase-Aided Amplification (RAA)-Exo-Probe and RAA-CRISPR/Cas12a Assays for Rapid Detection of Campylobacter jejuni in Food Samples. J Agric Food Chem 2022, 70 (30), 9557–9566.

(50) Kulesh, D. A.; Loveless, B. M.; Norwood, D.; Garrison, J.; Whitehouse, C. A.; Hartmann, C.; Mucker, E.; Miller, D.; Wasieloski, L. P., Jr.; Huggins, J.; et al. Monkeypox virus detection in rodents using real-time 3’-minor groove binder TaqMan assays on the Roche LightCycler. Lab Invest 2004, 84 (9), 1200–1208.

