## Supplementary material for "CRISPR/Cas12a-mediated ultrasensitive and on-site monkeypox viral testing": Supporting information.docx

**Expression and purification of cas12a**

The pMBP-LbCas12a was a gift from Jennifer Doudna (Addgene plasmid # 113431; http://n2t.net/addgene:113431; RRID:Addgene_113431). The plasmid pMBP-LbCas12a was transformed into *Escherichia coli* (*E. coli*) Rosetta (DE3) by heat shock. Totally 1 L aliquot of liquid Terrific-Broth medium with 100 μg/mL ampicillin was inoculated with 10 mL overnight cultures containing activated bacteria in the Luria-Bertani medium for the expansion of cultivation. Growth medium plus inoculant was grown at 220 rpm and 37 ℃ for 2 h and then at 200 rpm and 20 ℃ for another 20 min until the cell density at 600 nm (OD_600_) reached 0.6. Then, 0.5 mM isopropyl β-D-1-thiogalactopyranoside (IPTG) was added and the cells were cultured at 20 °C for 18 h. Cells were harvested by centrifugation, resuspended in lysis buffer (50 mM Tris-HCl, 500 mM NaCl, 0.25 mg/mL lysozyme, pH 8.0), and lysed by sonication. Protein purification included separation via high-affinity Ni-NTA resin (Genescript, China), MBP-digestion via TEV protease (Beyotime, China), and final acquisition via Dextrin Beads 6FF (Smart-Lifesciences, China). The purified LbCas12a was quantified by BCA protein determination reagent and stored at -80 °C.

**Sensitivity evaluation of PCR-Cas12a-MPXV fluorescence assay**

The PCR reaction system was carried out in a 10 μL solution containing 400 nM forward and reverse primers, 1 × Taq Master Mix, 1 μL serial 10-fold dilution (10^10-0^ copies/μL) of DNA templates, and ultrapure water. The temperature procedure of PCR was 95 °C for 3 min, 30 circles at 95 °C for 30 s, 55 °C for 30 s and 72 °C for 1 min, and final extension at 72 °C for 5 min. Then, the PCR-Cas12a-MPXV fluorescence assay was executed using the same condition as the RAA-Cas12a-MPXV fluorescence assay, except for PCR products in place of RAA products.

**Agarose gel electrophoresis**

For agarose gel electrophoresis, all of the reaction samples were incubated the same as the RAA-Cas12a-MPXV fluorescence assay procedures, and a 5 μL aliquot of the reaction solution was subsequently mixed with 1 μL of 6 × loading buffer (Sangon Biotech, China) containing 1 % GelRed (Biosharp, China). Then, agarose gel electrophoresis was performed with 3 % agarose gel at a constant voltage of 150 V for 30 min using 1 × TAE (40 mmol/L Tris-Acetate; 1 mmol/L EDTA; pH 8.0) as the running buffer, followed by imaging with an automatic digital gelatin image analysis system (Tanon, China).

**Table S1.** DNA sequences used in this study.

| **Name** | **Sequence (5’→3’) ^*^** |
| --- | --- |
| F1.1 | CAGCTCCAACGATACTCCTCCT |
| F1.2 | TCTACGACAATGGATGCTGATACACGGC |
| F2 | TACAGCTCCAACGATACTCCTCC |
| R1 | GACAGGGTTAACACCTTTCCAATAAAT |
| R2 | TTCCGTCAATGTCTACACAGGCA |
| crRNA1 | UAAUUUCUACUAAGUGUAGAUA*CAGUACUCAUUAAUAACG* |
| crRNA2 | UAAUUUCUACUAAGUGUAGAUA*UGAUGUUAUUCCGGUUAA* |
| CrRNA3 | UAAUUUCUACUAAGUGUAGAUA*CCGGAAUAACAUCAUCAA* |
| FQ reporter | FAM-TTATTT-BHQ1 |
| FB reporter | FAM-TTATTT-biotin |

* The italicized sequences in crRNAs are complementary to partial sequences of the target.

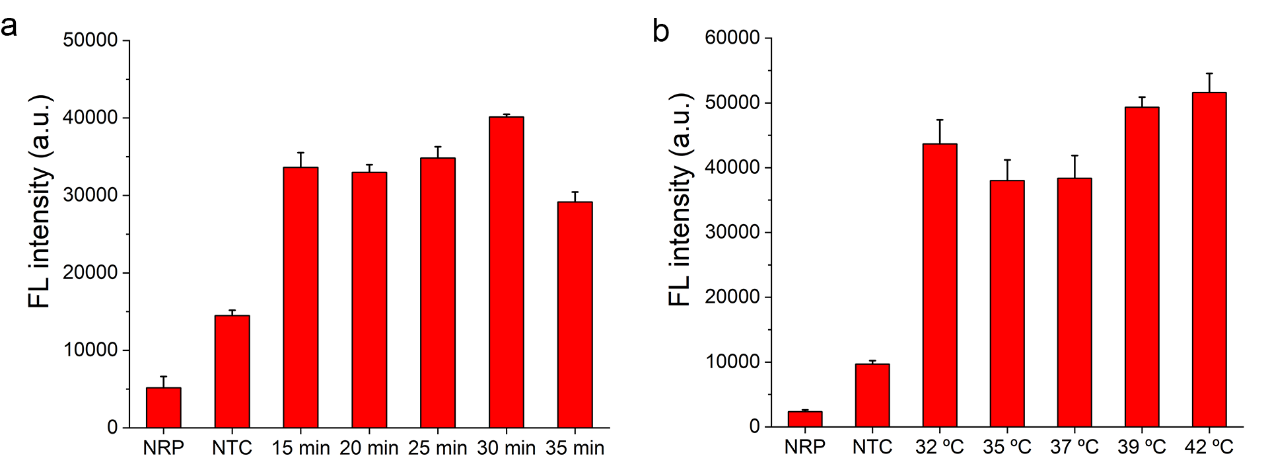

**Figure S1.** Optimization of RAA reaction conditions. (a) Fluorescence intensity of RAA-Cas12a-MPXV assay using RAA amplicons under different reaction times (15 min, 20 min, 25 min, 30 min, 35 min). (b) Fluorescence intensity of RAA-Cas12a-MPXV assay using RAA amplicons under different reaction temperatures (42 ℃, 39 ℃, 37 ℃, 35 ℃, 32 ℃). NRP, no RAA product. NTC, no template control.

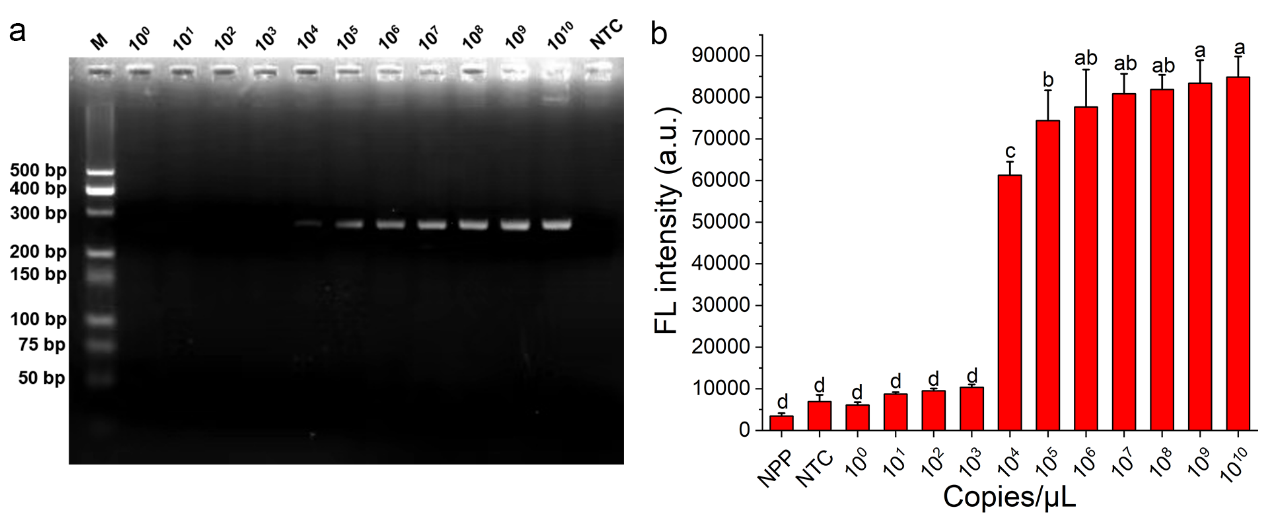

**Figure S2.** The evaluation of PCR sensitivity. (a) The agarose gel image of PCR products using serial 10-fold dilution (10^10-0^ copies/μL) of DNA templates. M, DNA marker. (b) Fluorescence intensity of CRISPR/Cas12a assay with PCR products using serial 10-fold dilution (10^10-0^ copies/μL) of DNA templates. NPP, no PCR product. NTC, no template control. Significance analysis results are labeled with different letters above the error bars (p < 0.05).
